# Targeted adaptive isolation strategy for Covid-19 pandemic

**DOI:** 10.1101/2020.03.23.20041897

**Authors:** Zoltan Neufeld, Hamid Khataee, Andras Czirok

## Abstract

We investigate the effects of social distancing in controlling the impact of the COVID-19 epidemic using a simple susceptible-infected-removed epidemic model. We show that an alternative or complementary approach based on targeted isolation of the vulnerable subpopulation may provide a more efficient and robust strategy at a lower economic and social cost within a shorter timeframe resulting in a collectively immune population.

## 1 SIR model

We consider the standard susceptible-infected-removed/recovered (SIR) epidemic model [1, 2] to represent the current COVID-19 pandemic:

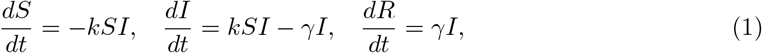

where the parameter *k* characterises the probability of transmission of infection from the infected (*I*) to the susceptible (*S*) fraction of the population, and *γ* is the rate of recovery, which is assumed to lead to immunity or death ((*R*)). The behavior of the model depends on a single non-dimensional parameter *R*_0_ = *k/γ* which is the number of new infections caused by a single infected in a fully susceptible population. The condition for an epidemic outbreak is *R*_0_ > 1, otherwise the infection dies out monotonously. Typical estimates of *R*_0_ for the COVID epidemic are roughly in the range 2 − 3.5 [3-5]. We will use *γ* = 0.1 day^− 1^ consistent with a typical recovery time of around 1 − 2 weeks [3-5]. A few example solutions of the model (1) for different values of *R*_0_ are shown in Fig. 1(a).

**Figure 1:**
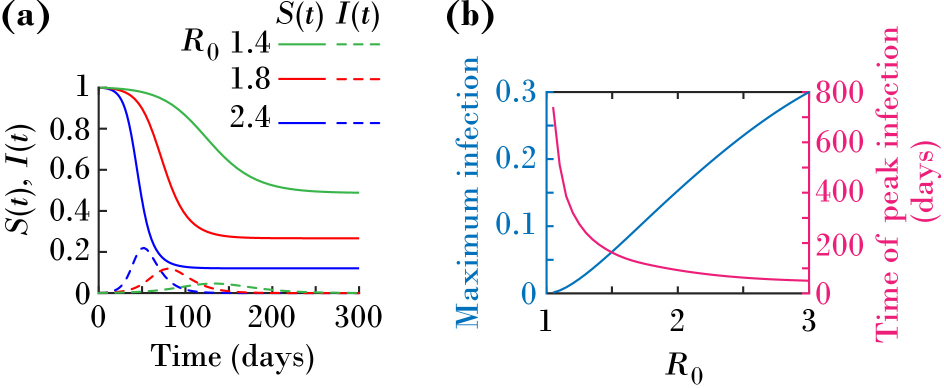
SIR model. (a) Solutions of the SIR model with *R*_0_ =1.4, 1.8, and 2.4. Solutions *S*(*t*) and *I*(*t*) represent the fractions of the susceptible and infected populations over time. Initial conditions: *S* = 0.9999, *I* = 0.0001, and *R* = 0. (b) Maximum fraction of infected individuals (left axis) and time at which the fraction of infected individuals has its peak (right axis) versus the basic reproduction ratio of infection, *R*_0_.

## 2 Social distancing

Since the COVID-19 infection results in mild or no symptoms in a large majority of the population, often may remain undetected which makes it difficult to prevent further transmission. The so called “social distancing” strategy aims to reduce social interactions within the population decreasing the probability of transmission of the infection, represented by the parameter *k* in the model. A strong reduction of social interactions may thus lead to *R*_0_ < 1 when the infection dies out, as *I*(*t*) ∼ exp(− *γ*(1 − *R*_0_)*t*). If the reduced *R*_0_ is still larger than unity, the result is a smaller epidemic outbreak, with lower total infected fraction of the population *I*_total_ = 1 − *S*(*∞*), and lower peak value of the infected fraction. The total infected population is the solution of the transcendental equation: ln(1 − *I*_total_) + *I*_total_ = 0. A side effect of this reduction, however, is that the duration of the epidemic outbreak increases significantly. The dependence of the peak infected fraction and the time to reach maximum infection are shown in Fig. 1(b). For example, starting with a reference value *R*_0_ = 2.4, the time to peak infection is around 65 days. Reducing *R*_0_ to 1.2, extends the time to peak infection to ∼ 320 days, while the maximum infected fraction is reduced from ∼ 22% to ∼ 1.5% and the total infected by the end of the epidemic decreases from ∼ 90% to ∼ 30%. Decreasing *R*_0_ by social distancing reduces the total infections and the peak infected fraction,which is critical due to limited hospital capacity, but it has the following possible drawbacks:

- if *R*_0_ remains > 1, the social distancing can significantly extend the duration of the epidemic, making it difficult to maintain the reduced transmission rate over a long time period in a large population.
- perhaps the most important problem is that there is no clear exit strategy until large scale vaccination becomes available. Since at the end of the epidemic a large proportion of the population remains susceptible to infection, after relaxation of social distancing, the population is highly susceptible to recurrent epidemic outbreaks potentially triggered by remaining undetected or newly imported infections from other regions/countries where the infection has not been eliminated yet.
- social distancing measures over extended period of time applied uniformly to a large population lead to widespread disruption of the functioning of the society and economy therefore it has a huge long term cost.

## 3 SIR model with vulnerable subpopulation

To address these issues, we consider an alternative or complementary strategy. An essential feature of the COVID-19 infections is that it produces relatively mild symptoms in the majority of the population, while it can also lead to serious respiratory problems mainly in the older population (> 70), or in individuals with pre-existing chronic diseases [6]. For example, the hospitalization rate of the symptomatic cases in the 20 − 29 age group is 1.2% out of which 5% requires critical care and 0.03% of the infections lead to death [7]. In contrast, for the 70 79 age group hospitalization rate is 24.3% out of which 43.2% is critical and the fatality ratio of the infected is 5.1%. Although the transition between these extremes is gradual, a fairly sharp transition takes place around the age of 65. In addition to age, pre-existing chronic diseases is an additional criteria for identifying a vulnerable group in the population. Based on the limited currently available data around 97 − 99% of deaths due to COVID-19 already had underlying chronic diseases, which is of course very common in the older age groups.

To take into account the markedly different age-dependent outcome of the COVID-19 infection, we extend the standard SIR model by separating the population into two compartments: the low-risk majority population with mild symptoms, and a vulnerable, mainly older population, where infection is likely to lead to hospitalization and death; see Fig. 2(a). Both sub-populations follow the SIR dynamics with the important difference that while in the majority population the infections are primarily caused by transmission within this sub-population from infected to susceptible; in the vulnerable minority the dominant route of new infections is via transmission from the majority population to susceptible vulnerable. We neglect transmission within the vulnerable population and the infection of low-risk susceptible by infected vulnerable; see Fig. 2(a), dashed transmissions.

**Figure 2:**
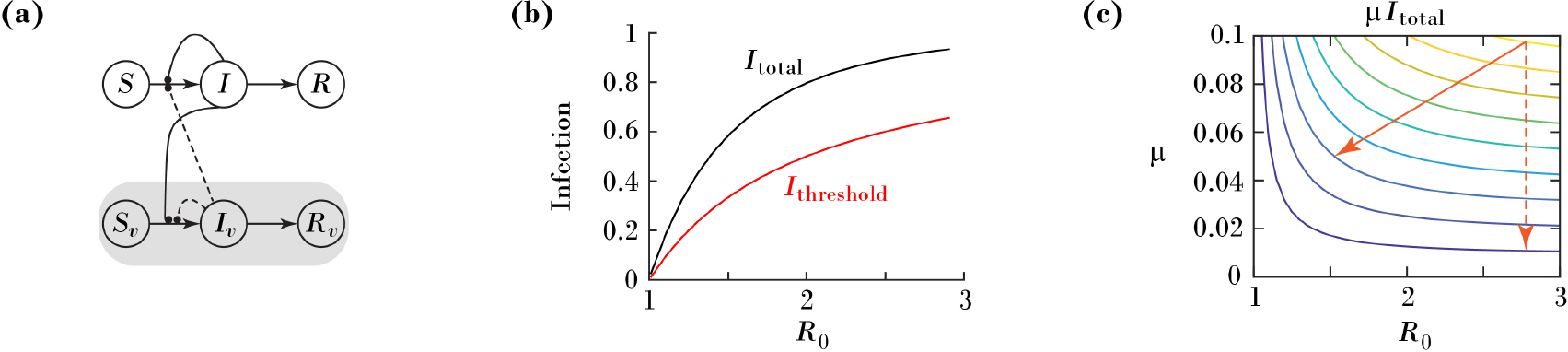
(a) Extended SIR model with vulnerable population. *S, I*, and *R* are proportions of susceptible, infected, and recovered individuals, respectively. Subscript *v* represents vulnerable individuals. Dashed curves: negligible transmissions of infection. (b) Black curve: total infected fraction *I*_total_ = 1 − *S*(*∞*) vs *R*_0_ in the SIR model. Red curve: 1 − 1*/R*_0_, minimum immune proportion needed for collective immunity. (c)Driver of infection in the vulnerable population: *µI*_*tot*_(*R*_0_) versus *µ* and *R*_0_. Diagonal solid arrow: social distancing. Dashed arrow: isolation.

The majority population follows the same SIR dynamics (1) as described above independently of the vulnerable population. The infection rate of the vulnerable population is described by

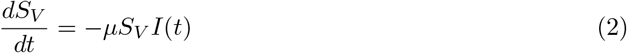

where *µ* represents the rate of transmission from the low-risk infected population (*I*) to vulnerable susceptible (*S*_*V*_). Thus, the relative proportion of newly infected vulnerable individuals (requiring hospitalization and possibility of death) per unit time is determined by the product *µI*(*t*).

We can also calculate the proportion of the vulnerable population infected over the whole course of the epidemic as

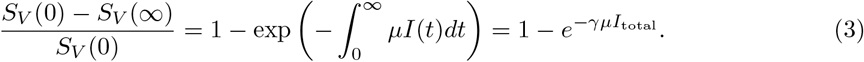

The total infected fraction of the low-risk population (i.e. standard SIR dynamics) at the end of the epidemic is shown in Fig. 2(b).

## 4 Targeted adaptive isolation

According to Equation (3), reducing the loss in *S*_*V*_ requires reducing the product *µI*_total_. While social distancing aims to reduce the total infected population by decreasing *R*_0_, this model helps to evaluate alternative or complementary measures targeting the reduction of parameter *µ* by isolating/shielding the vulnerable population from the infections. This could be achieved by clearly differentiated measures targeted to the high-risk group: restricting mobility, providing free home-delivery of food and medication, increased support addressing communication and healthcare needs, and providing separated living space where needed. Since the isolation strategy targets a sub-population, a radical isolation is likely to be more effective than uniform social distancing, and at a smaller cost for the economy and for the general functioning of the society.

Our model allows the comparison of general social distancing affecting both *k* and *µ* (represented by the diagonal arrow in Fig. 2(c)) and targeted isolation affecting *µ* only (vertical arrow in Fig. 2(c)). The overall fatality of the pandemic is primarily driven by the size of the infected vulnerable population, *µI*_total_. Depending on the epidemiological situation, the public response should be a mixture of the two efforts. For low values of *R*_0_, general social distancing is more effective and has the potential to suppress the epidemic. At higher *R*_0_ values, however, the optimal public response should focus more heavily on isolation of the vulnerable population. With limited resources available, when the strategy is primarily based on drastic targeted isolation over a shorter time, the end result is a collectively immune population resistant to further infections (“herd immunity”).

It is also apparent that the integral in Equation (3) can be reduced by decreasing *µ* in a time-dependent manner. This can be achieved by monitoring the progression of the infection *I*(*t*) via statistically representative testing of different regions and cities, and intensifying/relaxing the isolation of the vulnerable population accordingly.

Let us consider the case when the low-risk population is ∼75 − 80% of the total, and when infected 3% is hospitalized, with 0.2% requiring intensive care [7]. Assuming that the intensive care capacity is around 10-20 per 100000, this allows for ∼ 5% infected at the peak of the epidemic in the low risk population, which corresponds to *R*_0_ ∼1.4. By the end of the epidemic, this results in *I*_total_ ∼ 0.6 within the low-risk population so the immunity in the total population is 45%. Assuming that after the epidemic the transmission rate increases back to *R*_0_ = 2.4, the minimum immune fraction needed for herd immunity is around 60%. Therefore, relaxing social distancing should happen before the end of isolation allowing for a further increase of the immune proportion in the low-risk group to avoid the spread of infections in the high-risk population.

Our model and conclusions rely on the separation of the population based on age and health into two compartments with very different outcomes and applying differentiated protective measures while allowing for the development of immunity in the rest of the population.

At the time of writing, certain countries follow various combinations of social distancing and isolation. In Sweden, only moderate social distancing is implemented which will likely lead to development of immunity in the population. This strategy, however, without introducing targeted measures to protect the vulnerable population may lead to high mortality and over-saturation of the health care system.

Italy and Spain, on the other hand implemented severe social distancing, but so far this seems to be unable to stop the progress of the epidemic and may in fact be on track towards achieving large scale infection and immunity in the population. However, with too much focus on the implementation of uniform social distancing and no clear targeted measures for identifying and efficiently protecting the vulnerable sub-population can lead to a scenario with high mortality in spite of the high social and economic costs of an extended and potentially recurrent epidemic.

Another interesting observation is the striking difference between the mortality within the confirmed infected patients in Italy (∼10%) compared to Germany (∼1%). While there can be multiple reasons for this difference, it is possible that the closer social and family interactions between the older and younger generations in Italy, corresponds to a higher baseline value of *µ*, and/or the current exposure of the disease targets mainly the younger population in Germany with policies in place to decrease the exposure of the vulnerable population.

## Data Availability

N/A

